# Why has Nigeria’s neonatal mortality decline stalled? An ecological analysis of public health financing and macroeconomic instability, 1990–2024

**DOI:** 10.64898/2026.08.26.26361383

**Authors:** Obumneme Benaiah Ezeanosike, Edak Ezeanosike, Charity Ifeyinwa Anoke, Oluchi Okoro, Obinna Orjingene, Elizabeth Chukwu, Ugo Okoli

## Abstract

**Background:** Nigeria carries one of the world’s largest burdens of neonatal death and remains far from the Sustainable Development Goal target. Whether health financing and macroeconomic instability are associated with newborn survival has rarely been examined for neonatal mortality specifically.

**Methods:** We conducted an ecological time-series analysis of national annual data, covering 1990–2024 for macroeconomic models (n = 35) and 2000–2023 for health-financing models (n = 24), the periods for which published data exist; no values were imputed. Neonatal mortality came from the UN Inter-agency Group for Child Mortality Estimation 2025 round with 90% uncertainty intervals, and other series from the World Development Indicators. The primary model regressed log neonatal mortality on government health expenditure per capita (purchasing power parity), out-of-pocket share and currency instability, with a linear trend, a post-break trend spline and Newey–West standard errors; first differences without trend terms were the main sensitivity analysis. The break was located by segmented regression; currency instability was tested under four constructions.

**Results:** The decline broke around 2010, the trend moving from −0.74 to +0.14 deaths per 1,000 annually (F = 145.4, p < 0.001). The subsequent rise fell within estimation uncertainty (2012: 37.6, 90% interval 33.9–41.5; 2022: 39.3, 33.4–46.4), supporting stagnation rather than reversal; Demographic and Health Surveys concur, reporting 42 per 1,000 for the five years preceding the 1990 survey and 41 preceding the 2024 survey. Government health expenditure per capita was inversely associated with neonatal mortality (−0.040, 95% CI −0.051 to −0.029, p < 0.001; first differences −0.016, p = 0.033) and was the only expenditure measure surviving both specifications; share-of-GDP measures did not (p = 0.196 and 0.889) and correlated positively in raw terms. Currency instability showed no association under any construction (p = 0.65–0.83). Public expenditure per capita moved non-monotonically, peaking in 2005, falling by 2010 and recovering by 2023 to a level still below the 2005 peak.

**Conclusions:** Neonatal mortality in Nigeria is ecologically associated with public health expenditure per capita, but not with commonly used share-based measures, nor with currency instability. Rising public spending accompanied stalled progress, directing attention toward how health resources are converted into services. Annual modelled mortality estimates could not support year-to-year inference, a limitation relevant to comparable studies.

## Introduction

### The problem

Neonatal mortality has become the dominant component of child death worldwide, and Nigeria carries one of the largest national burdens [1]. The Sustainable Development Goal target 3.2.2 commits countries to reducing neonatal mortality to no more than 12 deaths per 1,000 live births by 2030 [2,3]. Nigeria remains far from that threshold. United Nations Inter-agency Group for Child Mortality Estimation figures place the national rate at 39.0 deaths per 1,000 live births in 2024, and the 2024 Nigeria Demographic and Health Survey reports 41 deaths per 1,000 for the five years preceding the survey [4].

What is striking is not only the level but the absence of movement. The same survey series recorded 42 deaths per 1,000 for the five years preceding its 1990 round, so more than three decades of health-system investment have produced almost no measurable change in newborn survival [4]. This stands in contrast to broader child survival, where under-five mortality fell from 132 to 110 deaths per 1,000 between the 2018 and 2024 survey rounds [4]. Nigeria is therefore saving the lives of infants and young children while making little progress for those in their first month.

The divergence is not accidental. Reductions in post-neonatal and child mortality have been driven substantially by interventions that are inexpensive, distributable and deliverable outside hospitals — insecticide-treated nets, oral rehydration, routine immunisation [5]. Newborn survival depends instead on facility-based capacity: skilled attendance at birth, functioning referral, thermal care, resuscitation equipment, oxygen delivery, injectable antibiotics and neonatal intensive care [6]. These are capital- and supply-intensive, and many of the required commodities are imported [2,7]. Newborn survival is consequently more sensitive to the financing and procurement capacity of the health system than are the gains achieved for older children.

Burdens within Nigeria are also markedly unequal, varying by geopolitical zone, by wealth quintile, and by whether birth occurs in a facility [4]. Barriers to skilled delivery care include distance, cost, insecurity and low maternal education [4,2].

### The knowledge gap

Nigeria finances health in a way that leaves the system unusually exposed. Total health expenditure stands at approximately 4.2% of gross domestic product, of which government sources account for only 14.3%, down from 18.3% in 2000. Out-of-pocket payment has averaged 71.5% of current health expenditure over the period examined here, exceeding 70% in most years. Government health spending has also remained close to 0.6% of GDP, far below the commitment made under the Abuja Declaration [8,9].

The macroeconomic environment surrounding that financing has been turbulent. Nigeria has experienced repeated oil price shocks, two recessions, sustained double-digit inflation, and successive exchange-rate regime changes, most recently the June 2023 liberalisation, after which the naira moved from an annual average of 426 per US dollar in 2022 to 1,479 in 2024 [10]. Where essential neonatal commodities are imported, currency depreciation transmits directly into procurement costs, and inflation erodes the real value of budgeted health allocations [10].

A substantial literature relates health expenditure to child mortality in low- and middle-income countries, but its findings are inconsistent [11,12]. Part of that inconsistency may be definitional. Studies variously examine total health expenditure, government expenditure, or out-of-pocket spending, and variously express these as shares of gross domestic product or in per capita terms, without establishing whether the choice is consequential [11]. A separate literature examines macroeconomic shocks and health outcomes [13,14], but the specific pathway from currency instability to newborn survival has rarely been tested directly [15,14].

Two further gaps motivate this study. First, most analyses treat child mortality as following a smooth monotonic decline, and few test whether the trend itself has changed — yet if progress has stalled, the timing of that change is the primary object of interest. Second, a large body of work regresses annual United Nations modelled mortality estimates on annual covariates without examining whether those estimates can support year-to-year inference [3,16]. Because such estimates are smoothed model outputs rather than measurements, this is not a minor technical matter.

### This study

This study examines Nigerian national annual data over 1990–2024 and asks four questions. First, has the decline in neonatal mortality changed in trend, and if so, when? Second, is government health expenditure associated with neonatal mortality, and does the answer depend on how expenditure is measured? Third, is currency instability associated with neonatal mortality? Fourth, can annual modelled mortality estimates support the year-to-year inference the literature routinely asks of them?

The contribution is fourfold. We locate and test a structural break in Nigeria’s neonatal mortality trend rather than assuming a constant rate of decline. We show that conclusions about health expenditure depend materially on whether spending is measured per capita or as a share of gross domestic product, which offers an explanation for the inconsistency of previous findings. We report a null result for currency instability that is robust across four constructions of the exposure. And we quantify the measurement properties of the modelled outcome series, establishing limits on what any annual analysis of these estimates can claim.

### Health expenditure and child survival

Whether public health spending improves child survival has been contested for three decades. The sceptical position was set out by Filmer and Pritchett, who found that roughly 95% of cross-national variation in child mortality was explained by per capita income, income distribution, female education, ethnic fragmentation and religion, leaving public health expenditure with little additional explanatory power [11,17]. Their estimated cost per child death averted through public spending ran to tens of thousands of dollars, against a small fraction of that for the interventions themselves, and they attributed the gap to how spending is allocated, to the net effect of additional public supply where private provision already exists, and to the efficacy of the public sector in converting appropriations into services [11].

Subsequent work using instrumental variable and dynamic panel methods has generally found stronger effects. Estimates of the elasticity of under-five mortality with respect to government health expenditure cluster between −0.25 and −0.42 [18], and analyses across low- and middle-income countries report elasticities of comparable magnitude for infant mortality, with the size of the effect conditional on the quality of governance [19]. A recent analysis spanning 188 countries between 2000 and 2019 examined health account indicators against neonatal and under-five mortality directly, stratified by income level [12].

Findings differ systematically by which component of expenditure is examined. Public expenditure has more often been found protective than total expenditure, and a dynamic panel of 37 African countries reports that the effect of public health expenditure on child mortality exceeds that of private expenditure once the two are entered jointly [20]. Evidence from sub-Saharan Africa specifically remains comparatively sparse [21]. This distinction matters most in settings where households finance the majority of health care, since total expenditure there largely reflects payments made at the point of illness rather than resources deployed in anticipation of it. Out-of-pocket expenditure has itself been associated with catastrophic financial burden and with delayed care-seeking [22,23], so an inverse association between out-of-pocket shares and mortality cannot be interpreted as beneficial.

Two further limitations of this literature bear on the present study. First, most analyses examine infant or under-five mortality rather than neonatal mortality, despite the two diverging substantially in both level and trend. Second, studies vary in whether expenditure is measured per capita, as a share of gross domestic product, or in nominal national currency, and rarely test whether that choice alters their conclusions. Nigerian studies illustrate the resulting inconsistency directly: published analyses report significant negative associations, significant positive associations, and no significant association at all, using broadly similar methods over broadly similar periods [24,25,26,27].

### Macroeconomic instability and neonatal care

That macroeconomic conditions affect child survival is well established. Using birth histories from 59 developing countries covering approximately 1.7 million births, Baird and colleagues estimated the elasticity of infant mortality with respect to per capita income at about −0.56, with a 1% decline in income associated with an increase in infant mortality of between 0.31% and 0.79%, and with larger effects for girls [15]. Africa-specific work has estimated the infant deaths attributable to the 2008–2009 financial crisis [28], and a global analysis of economic downturns between 1981 and 2010 reported associations with child mortality across country income groups [13]. A recent systematic review of economic growth and recessions in sub-Saharan Africa summarises this evidence for maternal and child health outcomes [14].

The evidence is not uniform, however. Effects have been found to be counter-cyclical in some middle-income settings, with mortality improving during contractions, echoing findings from high-income countries that mortality rises during economic expansions [29,30]. The direction and magnitude of the relationship therefore appear to depend on context, on the outcome examined, and on the period studied.

Currency instability is a distinct channel, and one with a specific mechanism in newborn care. Approximately 70% of medicines consumed in Nigeria are imported, most active pharmaceutical ingredients are imported even for locally finished products, and domestic manufacturing has operated well below capacity [7]. The national newborn plan identifies strengthening local production of neonatal commodities, including active pharmaceutical ingredients, as a priority action, and notes that facilities in remote or insecure areas lack reliable supply chains for essential newborn medicines [2]. Depreciation raises the local-currency cost of these inputs directly, and where budgets are appropriated in nominal terms their real purchasing power falls. Following the June 2023 liberalisation, medicine prices in Nigerian public and private pharmacies rose consistently over the subsequent year, attributed by the authors to import dependence and foreign exchange volatility, with stock-outs in public facilities expected to worsen as purchasing power declined [10].

The pathway extends beyond medicines to equipment. An assessment of oxygen access in secondary-level Nigerian hospitals found that of 57 oxygen concentrators tested, two were fit for use, and that fewer than a fifth of hypoxaemic children received oxygen [31]; a national case study identifies inadequate financing mechanisms and weak equipment maintenance among the principal constraints on oxygen security [32]. Facility assessments report correspondingly low levels of staff training in oxygen use and of oxygen administration at the point of care [33], and in the north-east armed conflict has damaged facilities and displaced health workers, further limiting access to skilled care at birth [34]. Since care of small and sick newborns depends on precisely such equipment and staffing, the mechanism linking currency movements to newborn survival is plausible on its face.

What is missing is a direct test. The macroeconomic literature summarised above examines income and output shocks; we located no study testing currency instability against neonatal mortality specifically. Given the scale of Nigeria’s recent exchange rate movements, that gap is worth closing, including the possibility that no association is detectable.

### Theoretical framework

The analysis is framed by the health production function derived from Grossman’s model of health capital [35]. In that framework health is a durable stock produced by combining medical care with time, education and other inputs, subject to budget constraints. Extended to the population level, the neonatal mortality rate can be treated as an output of a national health production process whose inputs include publicly financed health services, household resources and the real price of the commodities the system requires [36,37].

Three implications follow. Publicly financed inputs should be protective, because they substitute for household payment at the point of need. Movements in the real cost of imported inputs should reduce effective health production for a given nominal budget. And household income and the burden of out-of-pocket payment should govern whether care is sought in time [38]. Table 1 maps each variable to its theoretical channel and expected sign.

**Table 1.** Variables, theoretical channels and expected associations.

| Variable | Theoretical channel | Expected association with neonatal mortality | Basis |
| --- | --- | --- | --- |
| Government health expenditure per capita | Publicly financed inputs into health production; substitutes for household payment | Negative | Health production function |
| Government health expenditure, % of GDP | Fiscal priority accorded to health; denominator varies with output | Negative, but confounded by GDP movement | Health production function |
| Out-of-pocket share | Financial barrier at the point of care; delayed care-seeking | Positive | Demand for health under budget constraint |
| Currency instability | Real cost of imported neonatal commodities and equipment | Positive | Input price channel |
| Inflation | Erosion of real budget allocations and household purchasing power | Positive | Input price channel |
| GDP per capita growth | Household resources and fiscal capacity | Negative | Income effect |

**Table 2.**
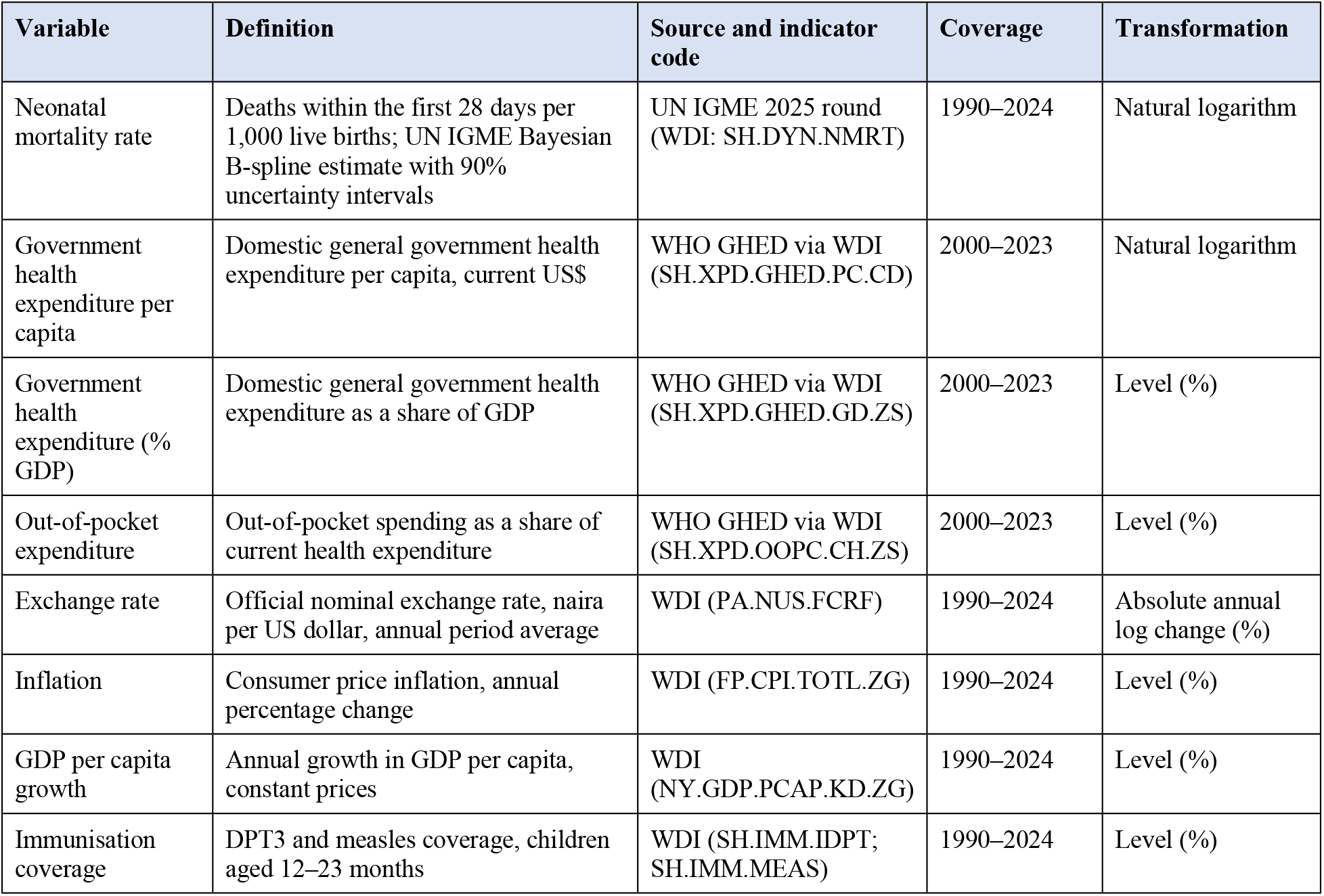
Variables, definitions and sources.

Because these expectations are stated in advance, the Discussion returns to each in turn and reports which are supported by the data and which are not.

## Methods

### Study design

This is an ecological time-series study of Nigeria using national annual data. The unit of analysis is the country-year. Because all variables are national aggregates, the analysis speaks to population-level association only and cannot support inference about individual newborns or households.

### Data sources and variables

All series were obtained from the World Bank World Development Indicators (WDI) [39] and, for neonatal mortality, from the United Nations Inter-agency Group for Child Mortality Estimation (UN IGME) [40]. Health-financing indicators originate from the WHO Global Health Expenditure Database and are compiled according to the System of Health Accounts 2011 framework [41,42]. Every series was downloaded in a single pass and mortality estimates are from the UN IGME 2025 estimation round. Because both UN IGME and the WHO Global Health Expenditure Database (GHED) re-estimate their entire back series at each release, series were downloaded complete, and the release vintage is reported so that results can be reproduced against a defined data version.

Government health expenditure was measured primarily in per capita US dollars rather than as a share of GDP. A share-of-GDP measure varies with its denominator, and Nigerian GDP moves substantially with oil prices, so the ratio can fall during an expansion and rise during a contraction while real resources are unchanged. The per capita measure is therefore closer to the resources actually available to the health system. Both measures are reported, and the difference between them is examined explicitly.

The exposure of interest was currency instability rather than the exchange rate level. The level measures cumulative depreciation and is dominated by trend; instability was therefore operationalised as the absolute annual log change in the official rate. Three alternative constructions — the signed log change, a rolling three-year standard deviation of the depreciation rate, and a rolling three-year coefficient of variation of the level — were carried through all analyses as sensitivity checks.

### Study period and analysis windows

Two windows were used, each determined by data availability rather than by preference. The macroeconomic model covers 1990–2024 (n = 35), the period over which UN IGME publishes neonatal mortality estimates for Nigeria. The health-financing model covers 2000–2023 (n = 24): WHO GHED begins in 2000 for all countries, and its most recent release extends to 2023, so no health-financing observation exists outside this window. The start of the health-financing window also coincides with the first full calendar year of civilian rule following Nigeria’s 1999 democratic transition.

### Missing data

No values were imputed, interpolated or extrapolated. Where a source publishes no estimate for a given year, that cell was left empty and the observation excluded from models requiring it. Consequently the health-financing models end in 2023 rather than 2024.

### Measurement properties of the outcome

UN IGME neonatal mortality estimates are the output of a Bayesian B-spline bias-adjusted model fitted to survey, census and registration data, not direct annual measurements [40,43,44]. Two consequences shape the analysis and are examined in the Results. First, the series is heavily smoothed: the standard deviation of its second differences is 0.030 of the standard deviation of its level, against 0.937 for consumer price inflation over the same period, and its first differences autocorrelate at 0.963. Year-to-year movement therefore reflects the estimation model as much as observed change. Second, precision deteriorates markedly toward the end of the series, the 90% uncertainty interval widening from ±3.7 deaths per 1,000 in 2010 (20% of the estimate) to ±8.1 in 2024 (41%). Uncertainty intervals were retained throughout and are reported alongside all point estimates.

As an external check, the modelled series was compared with the underlying survey observations that inform it, comprising Demographic and Health Surveys from 1990 to 2024, Multiple Indicator Cluster Surveys and the 2010 Malaria Indicator Survey, each plotted with its sampling standard error. Because retrospective birth histories are subject to recall error that increases with the length of the recall period, comparisons across surveys were restricted to the most recent period of each [45,46].

### Empirical strategy Series properties

Augmented Dickey–Fuller [47], Phillips–Perron [48] and Kwiatkowski–Phillips–Schmidt–Shin [49] tests were applied to each series in levels and first differences, with and without a deterministic trend. Log neonatal mortality did not reject the unit-root null under a constant-only specification (p = 0.131) but rejected it decisively once a broken deterministic trend was allowed (p = 0.0007), indicating that the apparent unit root reflects a structural break rather than genuine stochastic non-stationarity [50]. Modelling therefore proceeded on a broken-trend stationary basis, with cointegration-based specifications retained as sensitivity analyses rather than as the primary approach.

### Structural break identification

The break was located by segmented regression, fitting a two-segment linear spline for every candidate break year with at least six observations either side and selecting the year minimising the residual sum of squares. The break date was treated as estimated rather than assumed, and all subsequent models were re-estimated across the plausible range of candidate years as a sensitivity check; this approach follows the logic of tests for structural change with unknown break dates [51].

### Primary specification

The primary model regresses log neonatal mortality on the exposure and covariates together with a linear trend and a post-break trend spline:

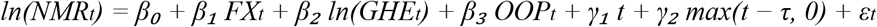

where τ is the estimated break year. Given the sample size, no specification carries more than three substantive regressors alongside the trend terms. Heteroskedasticity- and autocorrelation-consistent (Newey–West) standard errors with two lags are reported throughout [52]. Coefficients, standard errors, 95% confidence intervals, p-values and sample sizes are reported for every model; no model is summarised by direction and goodness of fit alone.

### Sensitivity analyses

Each result was re-estimated: in first differences without trend terms; across candidate break years; with and without trend and spline terms; using each of the four alternative instability measures; with the exposure entered at lags of zero, one and two years; weighting observations by the inverse variance implied by the UN IGME uncertainty intervals; and adding DPT3 and measles coverage as service-coverage controls. Autoregressive distributed lag models with Pesaran–Shin–Smith bounds testing [53] were estimated as a further check on whether conclusions depend on the treatment of non-stationarity. A finding was treated as robust only where sign and significance persisted across this set; results sensitive to specification are reported as such rather than at their most favourable.

Model adequacy was assessed by Breusch–Godfrey tests for serial correlation [54,55], Breusch–Pagan tests for heteroskedasticity [56], Jarque–Bera tests on residuals [57], Ramsey RESET for functional form [58], and variance inflation factors.

### Reporting

Reporting follows the Strengthening the Reporting of Observational Studies in Epidemiology (STROBE) statement, adapted for an ecological design in which the unit of analysis is the country-year rather than the individual. A completed checklist is provided as supplementary material, with notes indicating where items apply differently to aggregate data. Items concerning participants, exposure measurement at individual level, matching and loss to follow-up are not applicable and are marked as such.

### Software and reproducibility

Analyses were conducted in Python 3.12 using statsmodels 0.14 [59]. The cleaned dataset, the extraction script that retrieves each series from source with its release date, and the full analysis code are provided as supplementary material so that every table and figure in this manuscript can be regenerated from the original downloaded files without further download. Supplementary tables reporting the structural break search, the complete robustness set and the full unit root battery are also provided; these are summarised but not reproduced in full in the Results.

## Results

### Descriptive statistics

Table 3 summarises the analysis variables. Over 1990–2024 the neonatal mortality rate averaged 42.3 deaths per 1,000 live births, ranging from 49.8 to 37.6. The exchange rate is severely right-skewed (skewness 3.51), reflecting the June 2023 liberalisation: the annual average moved from 425.98 naira per US dollar in 2022 to 645.19 in 2023 and 1,478.97 in 2024. Absolute annual depreciation averaged 16.1% with a maximum of 144.0%. Government health expenditure averaged 27.2 PPP international dollars per capita and 0.64% of GDP, against total health expenditure of 3.67% of GDP, while out-of-pocket payment averaged 71.5% of current health expenditure, ranging from 60.2% to 77.4%.

**Table 3.** Descriptive statistics. JB p = Jarque–Bera test of normality.

| Variable | n | Mean | SD | Minimum | Median | Maximum | JB p |
| --- | --- | --- | --- | --- | --- | --- | --- |
| Neonatal mortality | 35 | 42.32 | 4.83 | 37.60 | 39.30 | 49.80 | 0.082 |
| Exchange rate (NGN/US\$) | 35 | 198.87 | 263.66 | 8.04 | 131.27 | 1478.97 | <0.001 |
| Absolute depreciation (%) | 34 | 16.06 | 28.84 | 0.00 | 5.97 | 143.96 | <0.001 |
| Inflation (%) | 35 | 18.71 | 15.87 | 5.39 | 13.01 | 72.84 | <0.001 |
| GDP per capita growth (%) | 35 | 1.43 | 3.94 | −8.34 | 1.92 | 12.21 | 0.543 |
| Govt health exp. p.c. (PPP \$) | 24 | 27.24 | 6.68 | 13.70 | 26.63 | 39.79 | 0.962 |
| Govt health exp. (% GDP) | 24 | 0.64 | 0.22 | 0.45 | 0.56 | 1.20 | 0.013 |
| Total health exp. (% GDP) | 24 | 3.67 | 0.59 | 2.49 | 3.58 | 5.05 | 0.673 |
| Out-of-pocket (% CHE) | 24 | 71.48 | 4.84 | 60.16 | 72.33 | 77.39 | 0.110 |

### Trends in the outcome

On the modelled series, neonatal mortality fell from 49.5 deaths per 1,000 live births in 1990 to 37.6 in 2012, then rose marginally to 39.0 by 2024. Segmented regression located the trend break at 2010, minimising the residual sum of squares; candidate years from 2009 to 2012 produced closely comparable fits, so the break is best characterised as occurring around 2010. The estimated trend was −0.737 deaths per 1,000 per year before the break and +0.142 after it. A segmented specification improved substantially on a single linear trend (F = 145.36, p < 0.000001).

The apparent post-break increase should not be interpreted as a reversal. The 2012 trough estimate of 37.6 carries a 90% uncertainty interval of 33.9 to 41.5, and the 2022 peak of 39.3 an interval of 33.4 to 46.4. These overlap almost entirely, and the 1.7-point rise represents 0.23 of the width of the 2012 interval. The data support a conclusion of stagnation, not of deterioration.

Figure 1 shows the four principal series in their native units. The pattern is corroborated by an independent measurement system, though the two do not agree throughout. Successive Nigeria Demographic and Health Surveys report 42 deaths per 1,000 for the five years preceding the 1990 survey and 41 for the five years preceding the 2024 survey, while under-five mortality fell from 132 to 110 per 1,000 between the 2018 and 2024 rounds. Survey figures refer to five-year recall windows rather than single calendar years, and are shown at the midpoint of each window in Figures 1 and 2. Progress in child survival more broadly has therefore continued without a corresponding improvement for newborns.

**Figure 1.**
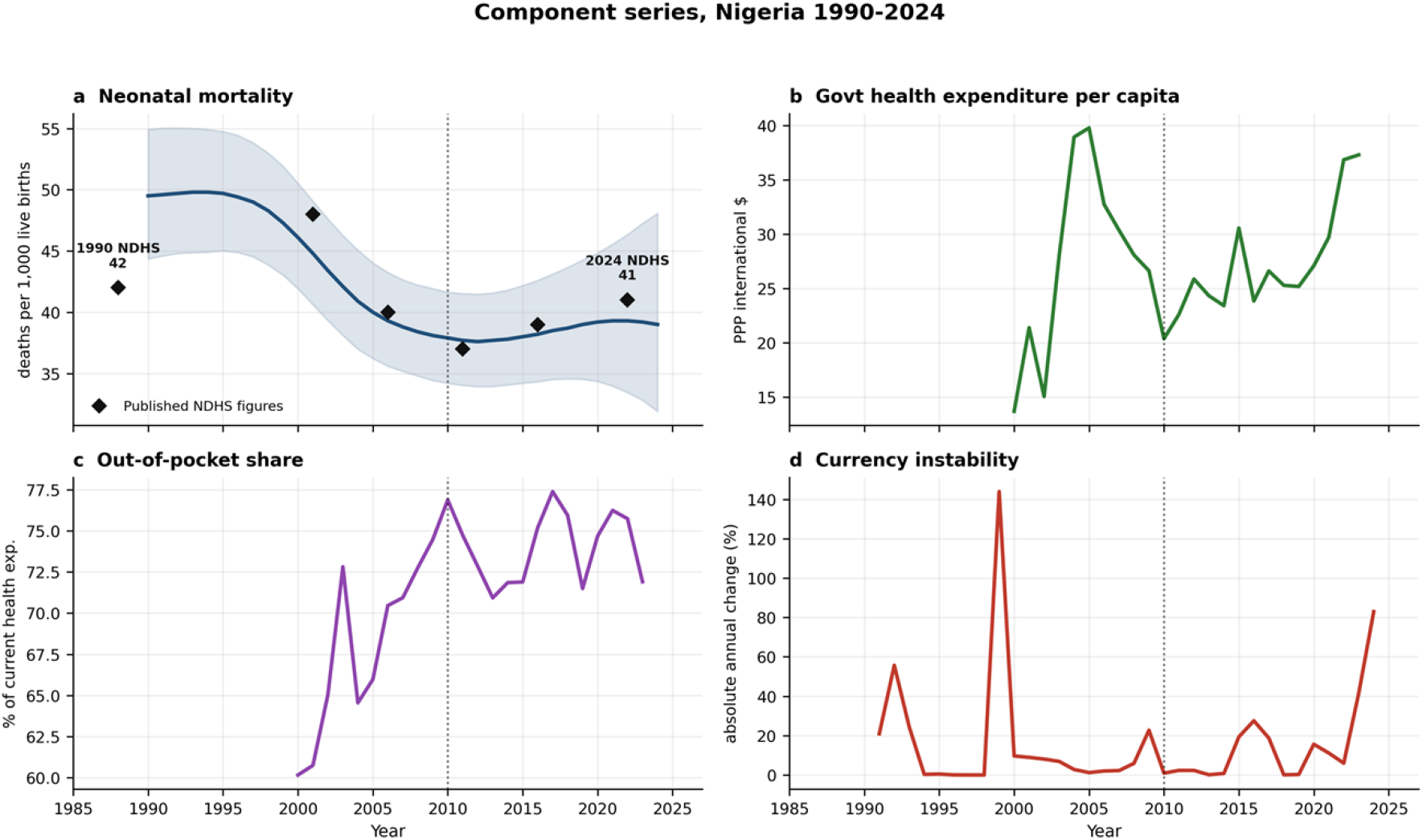
Component series in native units, 1990–2024. (a) Neonatal mortality per 1,000 live births, shaded band showing the 90% uncertainty interval; (b) government health expenditure per capita in purchasing power parity international dollars; (c) out-of-pocket payment as a share of current health expenditure; (d) currency instability, measured as the absolute annual log change in the official exchange rate. Health-financing series begin in 2000 and end in 2023, reflecting the coverage of the source database. Dotted line marks the estimated trend break in 2010.

**Figure 2.**
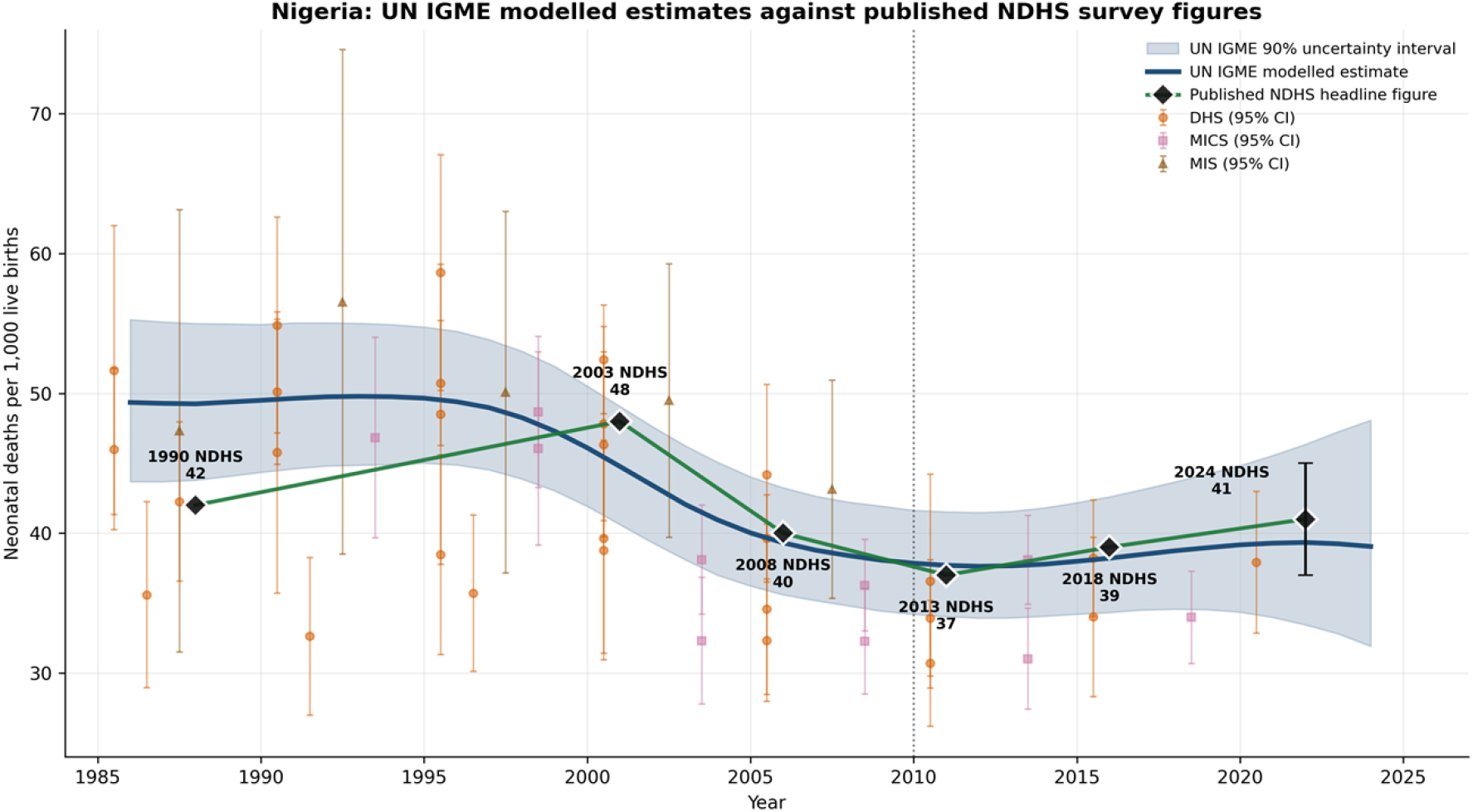
UN IGME modelled neonatal mortality estimates against published survey figures. The navy line and shaded band show the UN IGME modelled estimate with its 90% uncertainty interval. Black diamonds joined by the green line are the headline figures published in successive Nigeria Demographic and Health Survey reports (Figure 8.1 of the 2024 final report); each refers to the five years preceding that survey and is plotted at the midpoint of that recall window, so the 1990 survey figure of 42 per 1,000 appears at 1988 and the 2024 figure of 41 appears at 2022. The 2024 figure carries a 95% confidence interval of 37 to 45. Faint markers are the individual survey estimates held in the UN IGME database, shown with 95% confidence intervals derived from their sampling standard errors. Before the mid-2000s these estimates disagree by more than sampling error can explain and the modelled series diverges from the published figures; from the mid-2000s onward they converge and the two sources agree to within about one death per 1,000. DHS, Demographic and Health Survey; MICS, Multiple Indicator Cluster Survey; MIS, Malaria Indicator Survey.

### Measurement properties of the outcome

The modelled nature of the outcome constrains what can be inferred from it, and this was assessed directly. The standard deviation of the second differences of the neonatal mortality series is 0.030 of the standard deviation of its level; the corresponding ratio is 0.937 for consumer price inflation and 0.431 for the exchange rate. First differences of the mortality series autocorrelate at 0.963, against 0.171 for inflation.

Residual autocorrelation could not be removed by increasing model flexibility. Fitting polynomials in time of degree two, four and six produced R² of 0.901, 0.994 and 0.998 respectively, while residual first-order autocorrelation fell only from 0.941 to 0.797 and the Breusch–Godfrey test rejected at p < 0.001 throughout. This is a property of a smoothed series rather than a symptom of misspecification, and it means that year-to-year inference from these estimates is not well founded.

Estimate precision also deteriorates toward the present. The 90% uncertainty interval widened from ±3.7 deaths per 1,000 in 2010 (20% of the estimate) to ±5.2 in 2020 (27%) and ±8.1 in 2024 (41%). The years containing the currency float are therefore those in which the outcome is least precisely estimated.

### Time-series properties

Log neonatal mortality did not reject the unit-root null under a constant-only augmented Dickey–Fuller specification (p = 0.131), but rejected decisively once a deterministic trend was included (p < 0.001), and an ADF test on the residuals of a broken-trend regression rejected at p = 0.0007. The series is therefore stationary around a broken deterministic trend, and the apparent unit root reflects the structural break rather than a stochastic trend. Cointegration-based specifications were retained as sensitivity analyses but are not the appropriate primary framework.

Among the covariates, the depreciation measures and GDP per capita growth were stationary in levels, while inflation, government health expenditure and the out-of-pocket share were integrated of order one.

### Primary model

Table 4 reports the primary specification over 2000–2023 (n = 24). Government health expenditure per capita is negatively and significantly associated with neonatal mortality: a coefficient of −0.0398 (95% CI −0.0509 to −0.0286, p < 0.001) implies that a 10% increase in per capita public health spending is associated with approximately 0.4% lower neonatal mortality. Neither currency instability (p = 0.741) nor the out-of-pocket share (p = 0.978) is associated with the outcome. Both trend terms are highly significant, consistent with the break identified above.

**Table 4.** Primary model. Outcome is log neonatal mortality, 2000–2023, n = 24. Newey–West standard errors, two lags. R² = 0.987.

| Predictor | Coefficient | Std. error | 95% CI | p |
| --- | --- | --- | --- | --- |
| Absolute depreciation | 0.00004 | 0.00011 | −0.00018 to 0.00026 | 0.741 |
| ln(govt health exp. p.c., PPP) | −0.0398 | 0.0057 | −0.0509 to −0.0286 | <0.001 |
| Out-of-pocket share | −0.00001 | 0.00048 | −0.00095 to 0.00093 | 0.978 |
| Linear trend | −0.0185 | 0.0006 | −0.0198 to −0.0173 | <0.001 |
| Post-2010 trend spline | 0.0239 | 0.0009 | 0.0220 to 0.0257 | <0.001 |

Diagnostics were mixed and are reported in full. The model showed no evidence of residual serial correlation (Breusch–Godfrey p = 0.761), heteroskedasticity (Breusch–Pagan p = 0.118) or non-normality (Jarque–Bera p = 0.987), and R-squared was 0.987. The Ramsey RESET test, however, rejected at p = 0.002 for squared terms and p = 0.004 for cubic terms, indicating unmodelled curvature in the functional form.

This was investigated directly. Replacing the piecewise-linear trend with a quadratic trend resolved the problem (RESET p = 0.176) and left the expenditure coefficient essentially unchanged at −0.0369 (p < 0.001), against −0.0398 in the reported specification. The rejection therefore reflects the approximation of a smoothly curving series by two straight segments rather than instability in the estimated association, and is consistent with the smoothing properties of the outcome documented above. The piecewise specification is retained as primary because the break year is itself an object of interest; the quadratic-trend model is reported as a robustness check.

The first-difference model showed the complementary pattern, passing the RESET test (p = 0.308) and tests for heteroskedasticity (p = 0.900) and normality (p = 0.437) while rejecting on serial correlation (Breusch– Godfrey p < 0.001). No specification tested satisfied every diagnostic, which is the expected consequence of a smoothed outcome series and is examined further below.

The macroeconomic model over 1990–2024 (n = 35) produced no significant association for any macroeconomic variable once the break was accounted for: absolute depreciation p = 0.903, inflation p = 0.583 and GDP per capita growth p = 0.179, with both trend terms significant at p < 0.001.

### Which measure of health spending matters

The choice of expenditure measure proved consequential (Table 5). All four expenditure measures were significant in the break-adjusted specification, but only the two per capita measures retained significance in first differences. Government expenditure as a share of GDP (p = 0.196), total health expenditure as a share of GDP (p = 0.889) and the out-of-pocket share (p = 0.363) all failed.

**Table 5.**
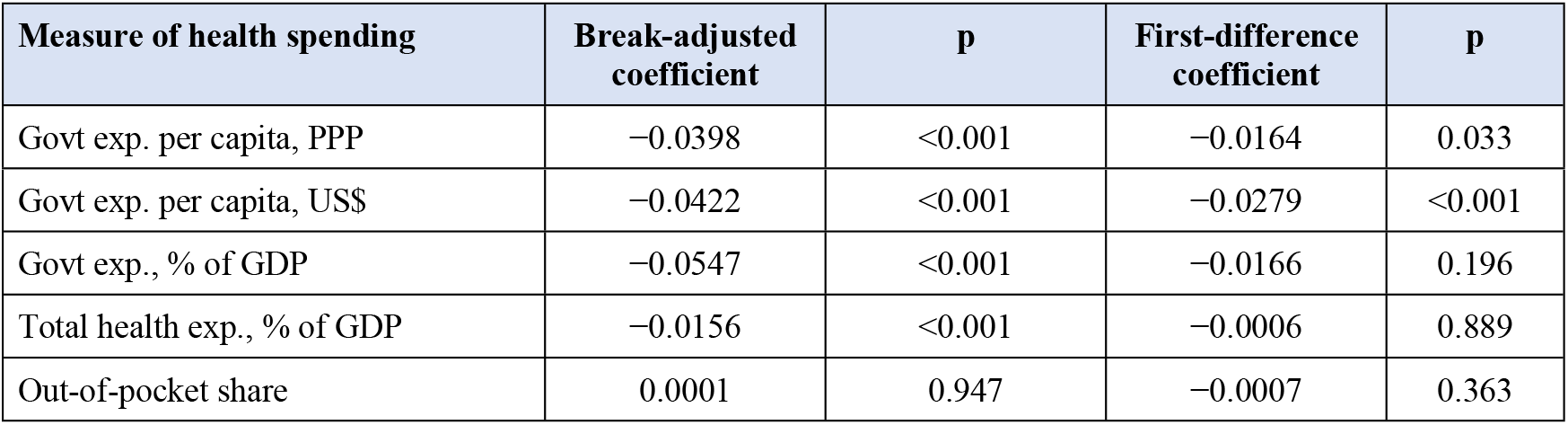
Association with log neonatal mortality by measure of health spending, 2000–2023. Each model contains the spending measure, the currency instability exposure and the out-of-pocket share, matching the covariate set of the primary model; break-adjusted models additionally contain the linear trend and post-break spline. Newey–West standard errors, two lags, n = 24 (23 in first differences).

| Measure of health spending | Break-adjusted coefficient | p | First-difference coefficient | p |
| --- | --- | --- | --- | --- |
| Govt exp. per capita, PPP | −0.0398 | <0.001 | −0.0164 | 0.033 |
| Govt exp. per capita, US\$ | −0.0422 | <0.001 | −0.0279 | <0.001 |
| Govt exp., % of GDP | −0.0547 | <0.001 | −0.0166 | 0.196 |
| Total health exp., % of GDP | −0.0156 | <0.001 | −0.0006 | 0.889 |
| Out-of-pocket share | 0.0001 | 0.947 | −0.0007 | 0.363 |

The failure of the share-of-GDP measures is interpretable. A ratio to GDP varies with its denominator, and Nigerian output moves substantially with oil prices, so the ratio may fall during an expansion and rise during a contraction while real resources are unchanged. The raw correlation between government expenditure as a share of GDP and log neonatal mortality is +0.380 — the opposite of the expected sign — against −0.425 for the PPP per capita measure. Expenditure per capita, which measures resources reaching the population, behaves as theory predicts; the ratio measures do not.

### Robustness of the expenditure association

The direction and magnitude of the association were stable throughout, but its statistical significance in differenced specifications depends on the covariate set and is reported here in full. In first differences the PPP-denominated coefficient ranged from −0.0144 to −0.0167 across covariate combinations, with p = 0.132 when entered alone, p = 0.112 with depreciation added, p = 0.038 with the out-of-pocket share added, p = 0.033 with both, and p = 0.028 with immunisation coverage additionally controlled. The coefficient is therefore stable while its precision improves as the specification is completed, but the differenced PPP estimate should be described as borderline rather than firmly established.

The US dollar denominated measure was significant across all differenced specifications (p = 0.0002 to 0.0013). Its stronger performance is expected, since that series embeds the exchange rate and therefore also captures general macroeconomic deterioration; for this reason the PPP measure is preferred as primary and the dollar measure is reported as sensitivity.

In levels without trend terms the PPP coefficient was −0.0494 (p = 0.0014). Across candidate break years from 2009 to 2013 the coefficient ranged from −0.0250 to −0.0593, all significant at p < 0.001; only a break imposed at 2008, outside the plausible range identified by the segmented regression, rendered it non-significant (p = 0.240). Weighting observations by the inverse variance implied by the uncertainty intervals left the estimate unchanged. In a direct comparison against the exchange rate in first differences the expenditure term held (p = 0.022) while depreciation did not (p = 0.092).

### Currency instability

No association was detected between currency instability and neonatal mortality under any construction of the exposure (Table 6), nor at lags of one and two years, where the cumulative three-year effect was 0.00005. This null holds with the float period included: the corrected series contains an 83.0% depreciation in 2024, so it does not reflect an absence of variation in the exposure.

**Table 6.**
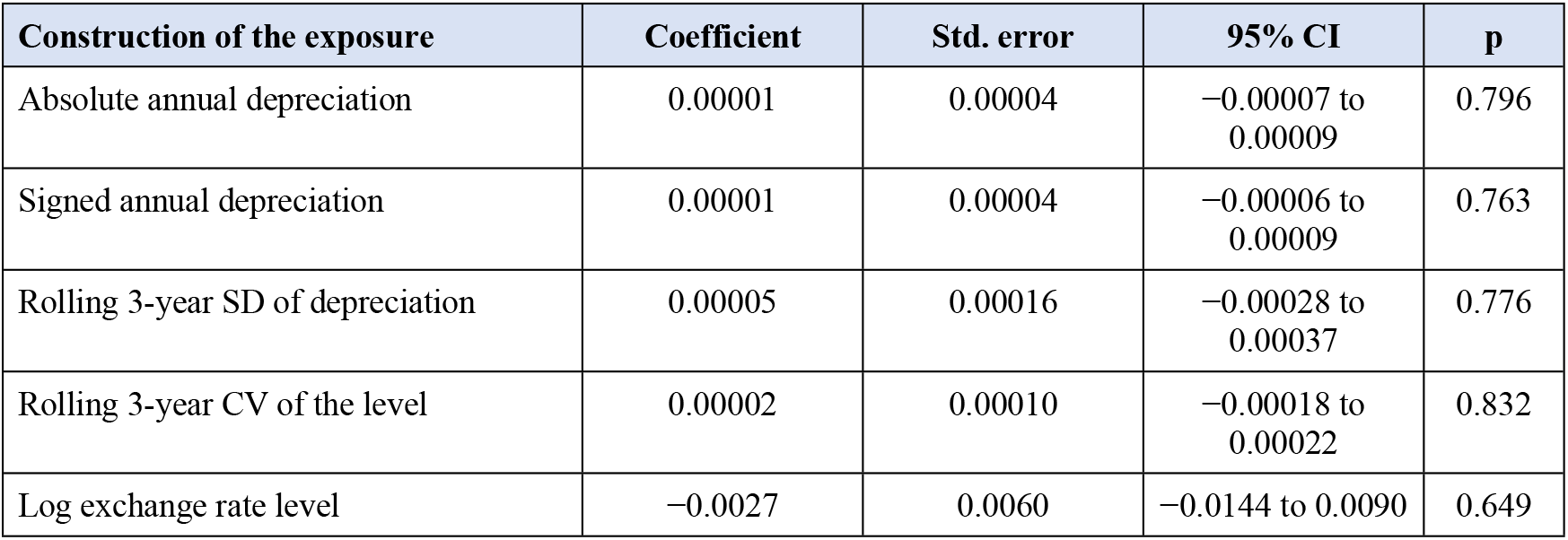
Currency instability under alternative constructions. First differences, 1990–2024, Newey–West standard errors.

Two qualifications apply. The official exchange rate was administratively managed for much of the study period, so it understates currency stress before 2023; a parallel-market premium would be a more sensitive measure but was not available for the full period. In addition, the float coincides with the years in which outcome precision is weakest, so this null partly reflects limited statistical power and should not be read as evidence of absence.

### Public spending and the stagnation

Government health expenditure per capita did not follow a simple upward path (Figure 3), and the pattern is more informative than an endpoint comparison suggests. It rose steeply from 13.70 PPP international dollars in 2000 to a peak of 39.79 in 2005, fell back to 20.38 by 2010, and then recovered to 37.30 by 2023. The 2023 value therefore remains 6.7% below the 2005 peak, and the 172% endpoint-to-endpoint increase over 2000 to 2023 reflects an unusually low base year rather than sustained growth. Over the post-break period from 2010 to 2023 the increase was 83%.

**Figure 3.**
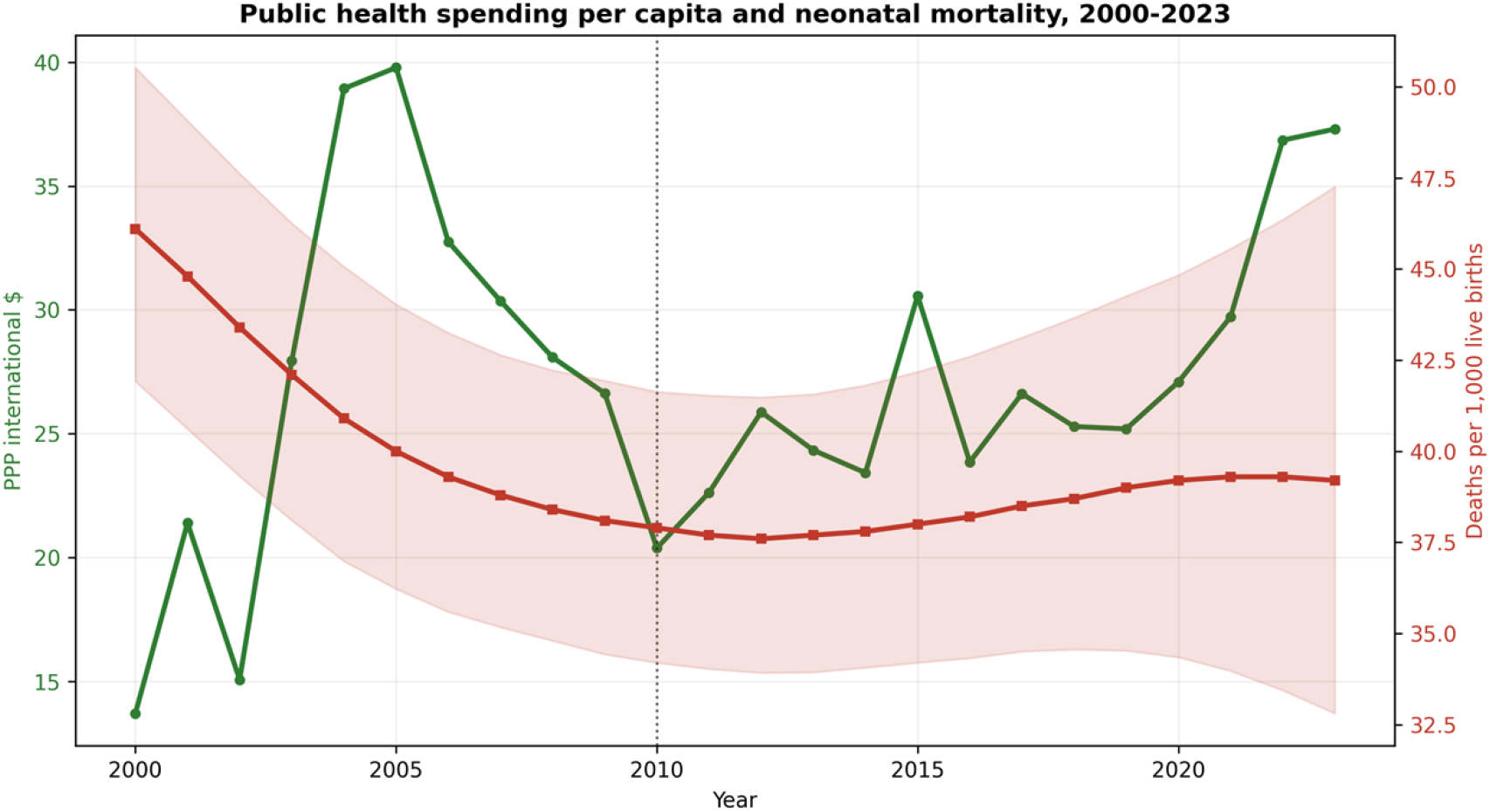
Government health expenditure per capita (purchasing power parity international dollars, left axis) and neonatal mortality with its 90% uncertainty interval (right axis), 2000–2023. Dotted line marks the estimated trend break in 2010.

**Figure 4.**
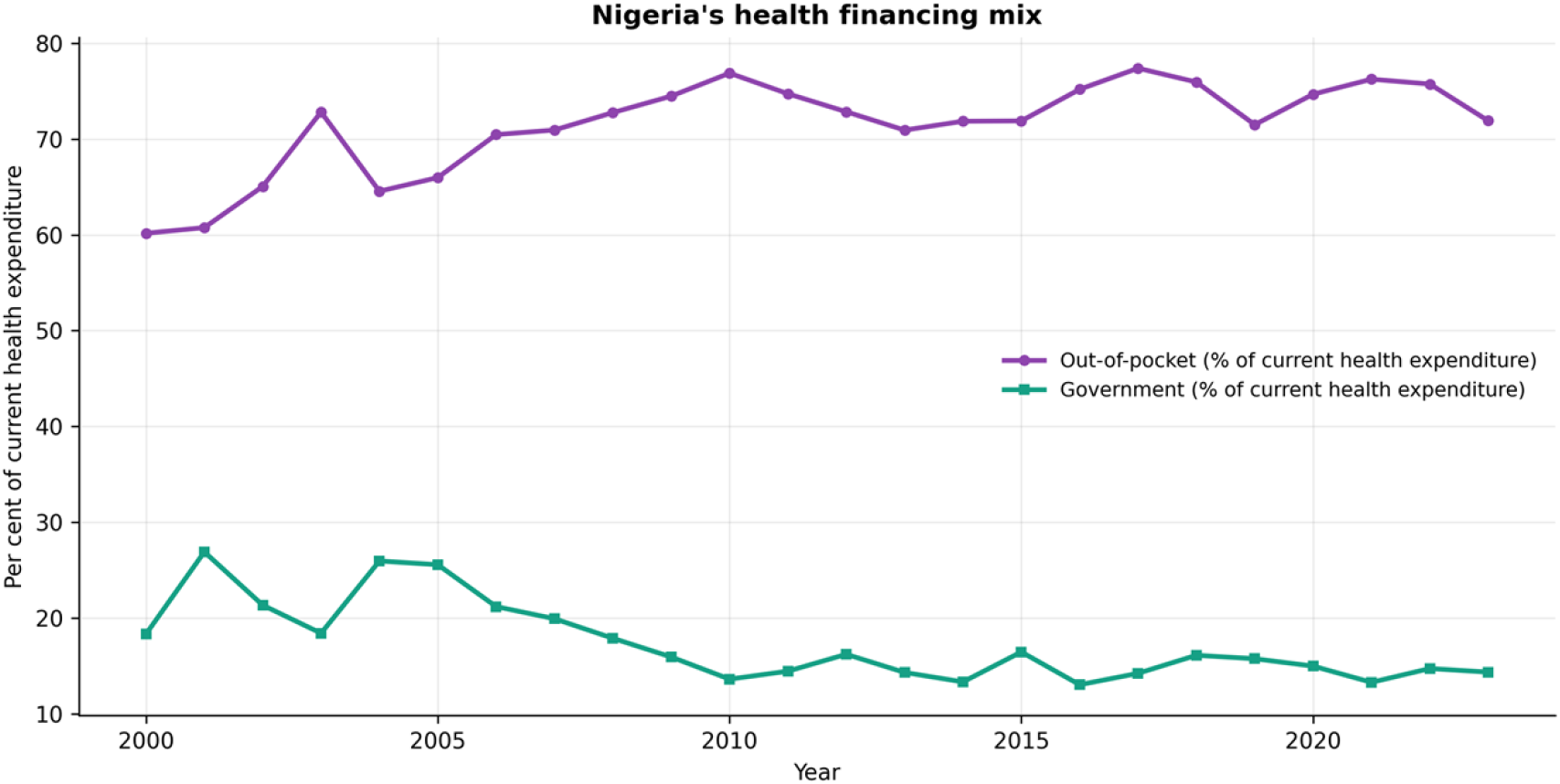
Composition of Nigerian health financing, 2000–2023: out-of-pocket payment and government spending as shares of current health expenditure.

The timing invites comment. The contraction in per capita public health spending between 2005 and 2010, a fall of 49%, coincides with the period in which the decline in neonatal mortality lost momentum, and the estimated trend break falls at the end of it. This is an observation about coincident timing in two series, not a causal finding, and the regression models already condition on trend; but it is consistent with the direction of the association reported above.

Over the same period the composition of health financing deteriorated. Total health expenditure remained close to 4% of GDP, but the government share of it fell from 18.3% in 2000 to 14.3% in 2023, having peaked at 25.6% in 2005, while out-of-pocket payment averaged 71.5% of current health expenditure, ranging from 60.2% in 2000 to a peak of 77.4% in 2017 and standing at 71.9% in 2023.

### Findings not interpreted

Three results are reported for completeness but are not interpreted, as each arises in specifications carrying seven parameters on 24 observations and none survives differencing. In the coverage-controlled model, measles coverage entered positively (p = 0.003) and DPT3 negatively (p = 0.009), a mutually contradictory pair that both became non-significant in first differences (p = 0.750 and p = 0.352). In the same specification the depreciation term reached p = 0.034 with a negative sign, contrary to the hypothesised direction, and was again null in differences (p = 0.598). In the comparison of spending measures, total health expenditure as a share of GDP entered positively (p = 0.043), most plausibly because the ratio rises as output contracts and therefore partly proxies recession.

### Summary

Nigeria’s decline in neonatal mortality stalled around 2010 and has not resumed, though the modest subsequent increase falls within estimation uncertainty and stagnation rather than reversal is the defensible conclusion. Government health expenditure per capita is negatively associated with neonatal mortality, robustly in level specifications and borderline in differenced ones, and is the only measure of health spending to survive the full robustness set. Currency instability and out-of-pocket expenditure show no association under any specification tested. Public health spending per capita nevertheless ended the period well above its 2000 level and 83% above its 2010 level without progress resuming, so the association identified here does not account for the stagnation itself.

## Discussion

### Principal findings

Four findings emerge. Nigeria’s decline in neonatal mortality stalled around 2010, the estimated trend moving from −0.74 to +0.14 deaths per 1,000 per year, though the subsequent increase falls within estimation uncertainty and stagnation rather than reversal is the defensible conclusion. Government health expenditure per capita is inversely associated with neonatal mortality, and is the only measure of health spending to survive the full robustness set. Currency instability shows no association under any construction of the exposure. And the annual modelled mortality series on which this and much comparable research depends cannot support the year-to-year inference routinely asked of it.

A fifth observation frames the others. Government health expenditure per capita did not rise steadily: it peaked in 2005, contracted by almost half to 2010, and recovered by 2023 to a level still below that peak. The association identified here is estimated from year-to-year variation conditional on trend and does not explain the stagnation.

### Interpreting the stagnation

The break identified here is not an artefact of the modelled outcome series. It is corroborated by an independent measurement system: successive Nigeria Demographic and Health Surveys report 42 deaths per 1,000 for the five years preceding the 1990 survey and 41 for the five years preceding the 2024 survey, while under-five mortality fell from 132 to 110 between the 2018 and 2024 rounds [4]. Two different approaches to measurement, one modelled and one survey-based, agree that newborn survival has not improved while child survival has.

This divergence is not unique to Nigeria. The Lancet Every Newborn Series established that reductions in neonatal mortality have been slower than for maternal and child mortality, slowest in the highest-burden countries and particularly in Africa [6], with under-five deaths halving over two decades while newborn progress lagged [5]. What Nigeria illustrates is that pattern in an extreme form, in a setting where the health-financing constraint is unusually severe.

The mechanism is plausible on the evidence. Gains in post-neonatal and child survival have been driven substantially by interventions that are inexpensive and deliverable outside facilities [5]. Newborn survival depends instead on care around the time of birth and on care of small and ill newborns, which together account for the largest share of avertable deaths, and the Series identifies the principal impediments to scaling that facility-based care as finance and workforce [6]. A country whose government finances 14% of its health expenditure is poorly positioned to scale precisely those services.

### Which theoretical expectations were supported

The health production function framework set out expected signs in advance [35][36,37]. Two expectations were met and three were not.

Publicly financed inputs were expected to be protective, and government health expenditure per capita was consistently negatively associated with neonatal mortality across every specification. The expectation of a positive association for out-of-pocket expenditure was not supported: the coefficient was indistinguishable from zero throughout. This is not evidence that financial barriers are unimportant, but rather that a national aggregate share of health spending is too blunt a measure of the barrier faced by any household, a limitation inherent to ecological data.

The input-price channel, by which currency depreciation was expected to raise the real cost of imported neonatal commodities, received no support. Nor did inflation or GDP per capita growth once the structural break was accounted for. The income channel in particular is well established in the wider literature [15,28], so its absence here requires explanation rather than acceptance, and we return to it below.

### Why the measurement of health expenditure matters

The most transferable finding of this study concerns measurement. Four candidate measures of health spending were significant in the break-adjusted specification, but only expenditure per capita survived differencing. Government expenditure as a share of gross domestic product, total health expenditure as a share of gross domestic product, and the out-of-pocket share all failed. More strikingly, the raw correlation between government expenditure as a share of GDP and log neonatal mortality is positive, the opposite of the theoretically expected sign, while the per capita measure in purchasing-power terms correlates negatively as predicted.

The explanation is arithmetic rather than substantive. A ratio to gross domestic product varies with its denominator, and Nigerian output moves substantially with oil prices, so the ratio can fall during an expansion and rise during a contraction while real resources are unchanged. A share-of-GDP measure is an indicator of fiscal priority; it is not a measure of the resources available to deliver care. Studies that use it are answering a different question from those that use per capita spending, and should not be expected to agree.

This offers a parsimonious explanation for the inconsistency of the wider literature [11,12][11]. It is also consistent with evidence that public expenditure is more often protective than total expenditure [20,12], since total expenditure in a setting where households finance three-quarters of health spending largely reflects payments made at the point of illness.

### Why rising expenditure has not restored progress

The central puzzle of this study is that substantial growth in real public health spending per capita has not restored progress in newborn survival. The trajectory is not a simple increase: spending peaked in 2005 at 39.79 PPP international dollars, contracted by 49% to 20.38 by 2010, and recovered to 37.30 by 2023, remaining 6.7% below its 2005 level. The 172% change measured between the endpoints of 2000 and 2023 reflects a low base year. Over the post-break period spending rose 83% without the decline in mortality resuming.

Three explanations have long been offered for the gap between the apparent potential of public health spending and its measured impact: how spending is allocated, the net effect of additional public supply, and the efficacy of the public sector in converting resources into services [11,17][11,60,61]. Nigerian fiscal evidence points clearly toward the third. Health sector allocations rose by 345% over five years while capital expenditure utilisation never exceeded 40.7%, with overall budget performance at 42% [60]. Resources were appropriated; less than half of the capital budget was converted into assets. Evaluations of the Basic Health Care Provision Fund, the principal vehicle for financing primary care, have similarly reported limited measurable effect on facility readiness and service utilisation [62]. On this evidence the binding constraint is absorptive capacity rather than allocation volume.

### Measured against Nigeria’s own targets

The scale of the shortfall is best seen against the country’s own commitments. The Nigeria Every Newborn Action Plan, adopted in 2016, set a baseline of 37 deaths per 1,000 from the 2013 Demographic and Health Survey and targets of 25 by 2020, 19 by 2025 and 15 by 2030, noting that the average annual rate of reduction between 2000 and 2015 had been 2.7% and that 5.2% would be required [2].

Neither the rate nor the targets were achieved. On the current estimation round the annual rate of reduction over 2000 to 2015 was 1.28%, less than half what the plan recorded, and over 2015 to 2024 it was −0.29%, that is, a slight increase. Table 7 sets the targets against what was achieved: neonatal mortality stood at 39.2 per 1,000 in 2020 against a target of 25, and at 39.0 in 2024 against a 2025 target of 19, more than double the target. Reaching 15 by 2030 from the 2024 position would require a reduction of about 14.7% per year, a rate Nigeria has not approached at any point in the observed record.

**Table 7.** Nigeria Every Newborn Action Plan targets for neonatal mortality against estimates achieved. Targets from the 2016 plan, baseline 37 per 1,000 (2013). Estimates and uncertainty intervals from the UN IGME 2025 round.

|  | NiENAP target | Estimate achieved | 90% uncertainty interval | Shortfall |
| --- | --- | --- | --- | --- |
| 2020 | 25 | 39.2 | 34.4 to 44.8 | +57% |
| 2024 (2025 target) | 19 | 39.0 | 31.9 to 48.1 | +105% |
| 2030 | 15 | — | — | requires 14.7% annual reduction |

What makes this more than a record of missed targets is that the plan itself identified the constraint. Its 2013 bottleneck analysis rated health financing as requiring major improvement, citing very low coverage of health financing schemes and the absence of any budget line for tracking maternal and newborn health resources, and its priority actions included not only allocating 15% of the government budget to health but also implementing a strategy to track expenditure, ensure prompt fund release and strengthen accountability [2]. The execution constraint identified here was named by the responsible ministry a decade ago. The evidence assembled above suggests it remains unresolved.

The composition of financing deteriorated over the same period. Total health expenditure remained close to 4% of gross domestic product, but the government share fell from 18.3% in 2000 to 14.3% in 2023, having peaked at 25.6% in 2005, while out-of-pocket payment averaged 71.5% of current health expenditure and exceeded 70% in most years [8,9]. Rising public spending per capita therefore occurred alongside a declining public share of a system that remained overwhelmingly privately financed at the point of use. Nigerian authors have already identified the efficiency of health spending as the under-explored question in this literature [24,25,26,27]; these findings suggest they are correct to do so.

### The absence of association with currency instability

No association was detected between currency instability and neonatal mortality under any of four constructions of the exposure, at any lag up to two years, or in any specification. This null is unusually well tested and holds with the 2023 liberalisation included, so it does not reflect an absence of variation in the exposure.

It should nonetheless be interpreted cautiously, for three reasons. First, the official exchange rate was administratively managed for much of the study period, so it understates the currency stress actually experienced; a parallel-market premium would be a more sensitive measure but consistent annual data were not available [63]. Second, the float coincides with the years in which outcome precision is weakest, the uncertainty interval reaching 41% of the point estimate by 2024. Third, and most importantly, the outcome series is smoothed in a way that removes precisely the short-run variation such a shock would produce.

That third point deserves emphasis because it bears on the wider literature. Studies that successfully detect income shocks in child mortality have generally used micro birth histories, constructing country series from observed deaths [15,28], not smoothed annual model estimates. The contrast suggests that failure to detect shock effects in modelled series may be a property of the data rather than of the world.

This does not mean the mechanism is absent. Nigerian evidence documents substantial increases in medicine prices following the 2023 devaluation, attributed to import dependence and foreign exchange volatility, with stock-outs in public facilities expected to worsen as purchasing power declined [10]. The pathway is well evidenced at the level of prices and availability. What these data cannot show is whether it registered in neonatal survival within the observation period.

### Comparison with previous Nigerian studies

Nigerian studies of health expenditure and child mortality have reached markedly divergent conclusions. Some report a significant negative association, some a significant positive association, and some no significant association at all [24,25,26,27]. These findings cannot all be correct, and the divergence is instructive.

Three features distinguish the present study. First, expenditure is measured in real per capita terms rather than as nominal aggregates or shares of output; the Results show this choice reverses the sign of the raw association. Second, a structural break is identified and accounted for; the earlier studies span periods of two to four decades while assuming a stable relationship throughout, and where a break is present, estimates that ignore it are not interpretable. Third, the outcome is neonatal rather than infant or under-five mortality, a distinction that matters given the divergence between them documented above.

The elasticity reported here is also considerably smaller than international estimates, which place the response of under-five mortality to government health expenditure between −0.25 and −0.42. Three factors plausibly account for the difference: the outcome here is neonatal rather than under-five mortality, and newborn survival responds to a narrower and more capital-intensive set of services; the design is a single-country time series rather than a cross-country panel, and therefore cannot exploit between-country variation in spending levels; and the inclusion of trend and break terms absorbs the long-run co-movement from which cross-sectional studies derive much of their identification. The estimate here should be read as a short-run within-country association, not as an elasticity comparable to cross-national work.

### What annual modelled mortality estimates can support

A methodological finding of this study extends beyond Nigeria. United Nations Inter-agency Group for Child Mortality Estimation figures are produced by a Bayesian spline regression model that, in the estimating agency’s own description, generates a smooth trend curve averaging over possibly disparate estimates from different data sources and extrapolating to a target year [40,43,44][3,16]. The consequences for regression analysis are measurable. The standard deviation of the second differences of the series is 0.030 of the standard deviation of its level, against 0.937 for consumer price inflation. First differences autocorrelate at 0.963. Residual autocorrelation could not be removed by any specification tested, persisting at 0.797 under a degree-six polynomial achieving an R-squared of 0.998.

Two implications follow. Standard inference on annual estimates of this kind is compromised in ways that are not addressed by conventional diagnostics, and a substantial body of published work regresses annual covariates on these series without examining whether they can bear such analysis. Reported findings from that literature, in either direction, warrant re-examination.

A second implication concerns unit root testing. Log neonatal mortality did not reject the unit-root null under a constant-only specification but rejected decisively once a broken trend was permitted. This is the well-established result that a structural break can produce the appearance of a unit root [50], and it means that cointegration-based frameworks, widely applied in this literature, may rest on a misdiagnosis of the data-generating process.

### Policy implications

Three implications follow from the findings, each tied to a specific result.

The finding that per capita expenditure is associated with neonatal mortality while shares of output are not suggests that monitoring frameworks anchored on budget shares may be poorly suited to tracking what reaches the health system. Real resources per person is the more informative quantity, and reporting it alongside share-based benchmarks would give a clearer picture of capacity.

The evidence that allocations rose sharply while capital utilisation remained below 41% suggests that, at current levels, the constraint on converting money into newborn survival lies at least as much in execution as in appropriation. Measures addressing procurement, release schedules and absorptive capacity may therefore yield more than further increases in nominal allocation alone [60][11,60,61].

The persistence of out-of-pocket payment at around 70% of health spending across a period of stagnating newborn survival, together with evidence that such payments push households into poverty [22,23], supports continued expansion of financial protection [64,65]. This recommendation rests on the equity and financial-protection literature rather than on any association detected in these data, where the out-of-pocket coefficient was null.

No recommendation follows from the exchange rate results, since no association was found.

### Strengths and future research

The principal strengths of this study are the treatment of measurement and the transparency of its robustness reporting. Every result is reported across the full specification set rather than at its most favourable, uncertainty intervals for the outcome are carried throughout, and the complete dataset and analysis code are available so that every table and figure can be regenerated from the original downloads.

The most promising extension is subnational. Neonatal mortality varies substantially across Nigerian states, from around 59 deaths per 1,000 in Kano to considerably lower rates in the South West [4], and health financing is substantially a state responsibility. Published subnational mortality estimates now exist [66,67], and a state panel with fixed effects would identify the relationship from within-state variation, removing the confounding that a national series cannot address. Given the limits established here for national annual data, that is where this question can most credibly be answered.

### Conclusion

Nigeria’s progress in reducing neonatal mortality stalled around 2010 and has not resumed. The modest subsequent increase falls within the uncertainty of the estimates, so stagnation rather than deterioration is the defensible conclusion; but three decades of survey evidence agree that newborn survival has not improved, even as child survival has.

Government health expenditure per capita is inversely associated with neonatal mortality, robustly in level specifications and more tentatively in differenced ones, and is the only measure of health spending tested here to survive that scrutiny. Measures expressed as shares of gross domestic product do not, and in raw terms carry the opposite sign, which offers an explanation for the inconsistency of previous findings. No association was detected between currency instability and neonatal mortality under any specification, though the outcome data are least reliable in exactly the years when currency movements were largest.

Public health spending per person nevertheless ended the period well above its 2000 level, and 83% above its 2010 level, without progress resuming. Whatever explains Nigeria’s stalled newborn survival, it is not simply the volume of public money, and the available fiscal evidence points toward the conversion of appropriations into functioning services. These are ecological associations and no causal claim is made; but they suggest that questions about how health resources are executed deserve at least the attention currently given to how much is allocated.

### Limitations

Several limitations constrain what can be concluded from this analysis, and they shape the tone of the conclusions that follow.

### The outcome is a modelled estimate rather than a measurement

This is the most consequential limitation. United Nations Inter-agency Group for Child Mortality Estimation figures are the output of a Bayesian model fitted to survey and census data, not annual observations. As reported in the Results, the series is heavily smoothed and its residual autocorrelation cannot be removed by any specification, however flexible. Year-to-year inference from these estimates is therefore not well founded, and the coefficients reported here should be read as summarising broad co-movement rather than annual response. The same caution applies to the substantial published literature that regresses annual covariates on these estimates without examining their properties.

A related constraint is that estimate precision deteriorates markedly in recent years, with the 90% uncertainty interval reaching 41% of the point estimate by 2024. The currency float of 2023–24 therefore falls in the period where the outcome is least reliably estimated. The null result for currency instability partly reflects this limitation and should not be interpreted as evidence that no such relationship exists.

### Design and inference

● The study is ecological. All variables are national aggregates, so no inference about individual newborns, mothers or households is possible, and associations observed at national level may not hold at individual level [68].
● The design is observational and the analysis correlational. No causal claim is made or supported. Reverse causation cannot be excluded, since deteriorating health outcomes may themselves influence budget allocation.
● National aggregation conceals substantial subnational variation. Neonatal mortality and health financing both differ markedly across Nigeria’s geopolitical zones and states, and a national series cannot detect whether associations hold within them.

### Sample size and statistical power

● The health-financing models rest on 24 annual observations and the macroeconomic models on 35. No specification carries more than three substantive regressors alongside trend terms, which constrains the covariates that can be included simultaneously.
● The association between government health expenditure per capita and neonatal mortality, while stable in magnitude, is statistically borderline in differenced specifications and its significance depends on the covariate set. It is reported as such rather than as firmly established.
● Null results for currency instability and out-of-pocket expenditure are consistent across specifications but cannot exclude effects too small to detect at this sample size.

### Measurement of the exposure

● Nigeria operated multiple or parallel exchange rates for much of the study period, and the official rate was administratively managed. The official series therefore understates currency stress before the 2023 liberalisation, when much of the adjustment occurred in parallel markets. A parallel-market premium would be a more sensitive exposure but consistent annual data were not available for the full period.
● Government health expenditure per capita expressed in current United States dollars mechanically incorporates the exchange rate. The purchasing-power-parity series is therefore used as the primary measure, with the dollar series reported only as sensitivity.

### Omitted determinants

Neonatal survival depends on determinants this analysis cannot include. Skilled birth attendance, antenatal care coverage, facility delivery rates, availability of neonatal intensive care, health workforce density, maternal education, poverty, insecurity and regional health-system inequality are all plausible contributors, and none is available as a complete annual national series for the study period. Immunisation coverage was included as a partial proxy for service-delivery capacity, but the coefficients obtained were mutually contradictory and did not survive differencing, so they should not be read as informative. The estimates reported here are therefore vulnerable to omitted-variable bias, most obviously if health-system capacity improved or deteriorated in ways correlated with public expenditure.

### Data vintage

Both the United Nations Inter-agency Group for Child Mortality Estimation and the WHO Global Health Expenditure Database re-estimate their entire back series at each release. The estimates reported here are specific to the 2025 estimation round and the July 2026 World Development Indicators vintage, and will change as those sources are revised. Health-financing data end in 2023 because no later observation is published; no value has been imputed to extend the series.

### What can and cannot be claimed

Taken together, these limitations support a narrow set of conclusions. The stalling of Nigeria’s neonatal mortality decline is robust, being visible in both modelled estimates and independent survey series. The apparent increase after 2012 falls within estimation uncertainty and cannot be described as a reversal. The association between public health expenditure per capita and neonatal mortality is consistent in direction across specifications but modest in size and borderline in the most demanding tests. The absence of association for currency instability and out-of-pocket expenditure is consistent but power-limited. None of these findings supports a causal interpretation.

## Declarations

### Abbreviations

ADF: Augmented Dickey–Fuller. ARDL: Autoregressive Distributed Lag. CHE: Current Health Expenditure. DHS: Demographic and Health Survey. GDP: Gross Domestic Product. GHED: Global Health Expenditure Database. HAC: Heteroskedasticity- and Autocorrelation-Consistent. KPSS: Kwiatkowski–Phillips–Schmidt–Shin. LMIC: Low- and Middle-Income Country. MICS: Multiple Indicator Cluster Survey. NDHS: Nigeria Demographic and Health Survey. NMR: Neonatal Mortality Rate. OOP: Out-of-Pocket. PPP: Purchasing Power Parity. SDG: Sustainable Development Goal. UN IGME: United Nations Inter-agency Group for Child Mortality Estimation. WDI: World Development Indicators. WHO: World Health Organization.

### Ethics approval and consent to participate

This study used only publicly available, aggregate, de-identified country-level secondary data from the World Bank World Development Indicators database and the United Nations Inter-agency Group for Child Mortality Estimation. It did not involve human participants, human biological material, identifiable personal data, clinical records, or any intervention. The study was therefore not submitted to a local ethics committee or institutional review board, and ethics approval was deemed unnecessary. No ethics reference number is applicable. Consent to participate was not applicable.

### Consent for publication

Not applicable. The manuscript contains no data from any individual person.

### Availability of data and materials

All data analysed in this study are publicly available. Macroeconomic and health-financing series were obtained from the World Bank World Development Indicators (https://data.worldbank.org) and neonatal mortality estimates with uncertainty intervals from the United Nations Inter-agency Group for Child Mortality Estimation (https://childmortality.org/data), with indicator codes, coverage and release dates given in Table 2 and in Supplementary File S9. The cleaned analysis dataset (S1), the complete analysis code (S5), an extraction script that retrieves each series from source (S6), and a full codebook (S8) are provided as supplementary material. Together these regenerate every table and figure reported here from the original downloaded files, without further download.

### Competing interests

The authors declare that they have no competing interests.

### Funding

This research received no specific grant from any funding agency in the public, commercial or not-for-profit sectors.

### Authors’ contributions

OBE conceived the study. OBE and CIA designed the analysis. OBE and CIA acquired and curated the data. OBE and EE conducted the formal analysis. OBE drafted the manuscript. OOk, OOr, EC and UO contributed to interpretation of the results. All authors critically revised the manuscript for important intellectual content, approved the final version, and agree to be accountable for all aspects of the work.

### Use of artificial intelligence

The authors used a generative artificial intelligence assistant (Claude, Anthropic) during the final preparation of this manuscript to support extraction of publicly available data and code development, and editing of text. The tool was not used to generate data or to generate references. All source data were downloaded directly from the cited public repositories, all analyses are reproducible from the supplied code, and all references were retrieved and checked by the authors against the original publications. The authors reviewed and verified all content and take full responsibility for the accuracy and integrity of the work.

## Data Availability

All data analysed in this study are publicly available. Macroeconomic and health-financing series were obtained from the World Bank World Development Indicators (https://data.worldbank.org) and neonatal mortality estimates with uncertainty intervals from the United Nations Inter-agency Group for Child Mortality Estimation (https://childmortality.org/data),

## Notes

### Competing Interest Statement

The authors have declared no competing interest.

